# Persistent SARS-CoV-2 infection and intra-host evolution in association with advanced HIV infection

**DOI:** 10.1101/2021.06.03.21258228

**Authors:** F Karim, MYS Moosa, BI Gosnell, S Cele, J Giandhari, S Pillay, H Tegally, E Wilkinson, JE San, N Msomi, K Mlisana, K Khan, M Bernstein, N Manickchund, L Singh, U Ramphal, COMMIT-KZN Team, W Hanekom, RJ Lessells, A Sigal, T de Oliveira

**Affiliations:** Africa Health Research Institute, Durban, South Africa; School of Laboratory Medicine and Medical Sciences, University of KwaZulu-Natal, Durban, South Africa; Department of Infectious Diseases, University of KwaZulu-Natal, Durban, South Africa; KwaZulu-Natal Research Innovation & Sequencing Platform, School of Laboratory Medicine and Medical Sciences, University of KwaZulu-Natal, Durban, South Africa; Discipline of Virology, University of KwaZulu-Natal, School of Laboratory Medicine and Medical Sciences and National Health Laboratory Service, Durban, South Africa; National Health Laboratory Service, Johannesburg, South Africa; Centre for the AIDS Programme of Research in South Africa (CAPRISA), University of KwaZulu-Natal, Durban, South Africa; Max Planck Institute for Infection Biology, Berlin, Germany; Department of Global Health, University of Washington, Seattle, United States

## Abstract

While most people effectively clear SARS-CoV-2, there are several reports of prolonged infection in immunosuppressed individuals. Here we present a case of prolonged infection of greater than 6 months with shedding of high titter SARS-CoV-2 in an individual with advanced HIV and antiretroviral treatment failure. Through whole genome sequencing at multiple time-points, we demonstrate the early emergence of the E484K substitution associated with escape from neutralizing antibodies, followed by other escape mutations and the N501Y substitution found in most variants of concern. This provides support to the hypothesis of intra-host evolution as one mechanism for the emergence of SARS-CoV-2 variants with immune evasion properties.

## Introduction

Coronavirus disease 2019 (COVID-19) is an acute respiratory illness caused by the severe acute respiratory syndrome-related coronavirus 2 (SARS-CoV-2). While most people effectively clear SARS-CoV-2 infection^1,2^, there are now several reports of prolonged infection associated with underlying immunosuppression^3-14^. In many of these cases, there has been evidence of intra-host evolution, characterised by the emergence of viruses with mutations in the spike glycoprotein. It has been hypothesized that intra-host evolution may be one mechanism for the emergence of SARS-CoV-2 variants^4,15,16^. Here we describe a case of prolonged SARS-CoV-2 infection in a person with advanced Human Immunodeficiency Virus (HIV) and antiretroviral treatment failure without either clinically induced immunosuppression or neutralizing antibody based treatment for SARS-CoV-2 infection, and present analysis of the virus evolution over time.

### Case Report

Am HIV-positive female in her late 30s was admitted to hospital in September 2020 with sore throat, cough and dyspnoea. Symptom onset was 12 days prior to admission. She had been on HIV antiretroviral therapy (ART) since 2006, most recently a fixed-dose combination of tenofovir, emtricitabine and efavirenz. Other relevant medical history included a single episode of tuberculosis in 2006, and chronic asthma for which she received inhaled budesonide and salbutamol. On the day of admission (day 0), SARS-CoV-2 reverse-transcriptase polymerase chain reaction (RT-PCR) of a nasopharyngeal swab specimen was positive (Allplex™ 2019-nCoV Assay, Seegene Inc., Seoul, South Korea; mean cycle threshold (Ct) 18.5). Sputum Xpert® MTB/RIF Ultra (Cepheid, Sunnyvale, CA) was negative. Other routine investigations are shown in Table S1. She was managed in a general COVID-19 ward, received oxygen via face mask and a six-day course of dexamethasone, and was discharged home after nine days.

During the hospital admission, she was enrolled in a prospective cohort study exploring the effect of HIV infection on the natural history and immune response to SARS-CoV-2 infection^17^. She was reviewed at enrolment (day 6 post-admission and day 16 post-symptom onset) when she was placed on non-high flow supplemental oxygen, and in the clinic on day 20 and day 34, when she was asymptomatic. On day 71, she complained of chest tightness. Oxygen saturation (SpO_2_) was documented to drop from 96% to 76% on exertion. Chest X-ray showed non-specific perihilar infiltrates. She was treated empirically for *Pneumocystis jiroveci* pneumonia on an ambulatory basis with 21 days of co-trimoxazole and prednisone. She was reviewed on day 106 when she reported only fatigue, and on day 190 when she was asymptomatic. On day 190, antiretroviral therapy was switched to a fixed-dose combination of tenofovir, lamivudine and dolutegravir. On day 206, she achieved HIV viral load suppression (i.e. < 50 cp/ml).

## Methods

### Ethical considerations

The protocol for the prospective cohort study exploring the effect of HIV and TB on the natural history and immune response to SARS-CoV-2 infection was approved by the Biomedical Research Ethics Committee of the University of KwaZulu-Natal (ref. BREC/00001275/2020) and the KwaZulu-Natal Provincial Health Research and Ethics Committee (ref. KZ_202005_003). We obtained written informed consent from all participants. Additionally, we obtained written informed consent for publication from the participant described in this report.

### Specimen collection and laboratory testing

We collected nasopharyngeal swabs and blood specimens on day 6 (day of study enrolment), day 20, day 34, day 71, day 106, day 190, day 204, day 216 and day 233. We performed SARS-CoV-2 RT-PCR testing on all swabs using either the Taqpath™ COVID-19 RT-PCR assay (Thermo Fisher Scientific, Waltham, MA) or the Allplex™ 2019-nCoV Assay (Seegene Inc., Seoul, South Korea). Anti-SARS-CoV-2 antibodies in the blood were measured using the Test-it CoV-2 IgM/IgG test kit (Lifeassay Diagnostics, Cape Town, South Africa), which detects IgM and IgG against the S1 subunit of spike. HIV-1 viral load was measured using the Abbott *m*2000 RealTi*m*e assay (Abbott Laboratories, Abbott Park, IL) and CD4+ count using the BD FACSCanto™ (BD Biosciences, San Jose, CA). We performed antiretroviral drug level testing using liquid chromatography coupled to tandem mass spectrometry (LC-MS/MS). Sample analysis was performed using an Agilent High Pressure Liquid Chromatography (HPLC) system (Agilent Technologies, Santa Clara, CA) coupled to the AB Sciex 5500 (AB Sciex, Framingham, MA), triple quadrupole mass spectrometer equipped with an electrospray ionization (ESI) TurboIonSpray source. The LC-MS/MS method was developed and optimised for the quantitation of tenofovir, emtricitabine, efavirenz, lopinavir, ritonavir, nevirapine, zidovudine, lamivudine, abacavir, atazanvir and dolutegravir in the same sample. We performed HIV-1 sequencing on all specimens with viral load >1000 copies/mL using the Illumina MiSeq instrument (Illumina, Inc., San Diego, CA) and used the Stanford HIV drug resistance database (version 9.0) for genotypic resistance interpretation (Supplementary Appendix).

### SARS-CoV-2 whole genome sequencing

Six specimens collected as part of the cohort study (day 6 – day 190) and the residual nasopharyngeal swab specimen collected for routine diagnosis on day 0 were all processed at the KwaZulu-Natal Research Innovation & Sequencing Platform laboratory (KRISP). We extracted SARS-CoV-2 ribonucleic acid (RNA) using the Viral NA/gDNA Kit on the automated chemagic™ 360 system (PerkinElmer, Inc., Waltham, MA). We converted RNA to cDNA using the Superscript IV First Strand synthesis system (Life Technologies, Carlsbad, CA) and random hexamer primers, using the ARTIC V3 protocol^18^. We performed SARS-CoV-2 whole genome amplification by multiplex PCR using primers designed on Primal Scheme (http://primal.zibraproject.org/), generating 400bp amplicons with an overlap of 70bp to cover the 30kb SARS-CoV-2 genome. We cleaned up PCR products using AmpureXP purification beads (Beckman Coulter, High Wycombe, UK) and quantified the products using the Qubit dsDNA High Sensitivity assay on the Qubit 4.0 instrument (Life Technologies, Carlsbad, CA). We then used the Illumina® Nextera Flex DNA Library Prep kit (Illimuna, Inc., San Diego, CA) to prepare indexed paired end libraries of genomic DNA. We normalized sequencing libraries to 4nM, then pooled and denatured with 0.2N sodium acetate. We spiked the 12pM sample library with 1% PhiX (PhiX Control v3 adapter-ligated library used as a control). We sequenced libraries on a 500-cycle v2 MiSeq Reagent Kit on the Illumina MiSeq instrument (Illumina, Inc., San Diego, CA). Full details of our amplification and sequencing protocol have been previously published^19,20^. We assembled paired-end fastq reads using Genome Detective 1.132 (https://www.genomedetective.com) and the Coronavirus Typing Tool^21^. Low-quality mutations were filtered out of the initial assembly generated from Genome Detective using bcftools 1.7-2 mpileup method. All mutations were confirmed visually with BAM files using Geneious software (Biomatters, Auckland, New Zealand). For analysis of low frequency mutations, we used LoFreq v.2.1.5^22^ to call intrahost variants relative to the Wuhan-Hu-1 NC_045512.2 reference. We employed a false discovery rate (FDR) cut-off of 1%. Additionally, LoFreq automatically applies default filters including minimum sequencing depth of x10, dynamic Bonferroni correction for variant quality and strand bias filtering. We annotated the variants using snpEff v4.5^23^ and extracted them for additional processing and visualizing using SnpSift v.4.3t^23^.

Genomes in this study were analyzed against a global reference dataset of 3566 genomes, including 486 genomes from South Africa, using a custom build of the SARS-CoV-2 NextStrain (https://github.com/nextstrain/ncov). The workflow performs alignment of genomes, phylogenetic tree inference, tree dating and ancestral state construction and annotation. The phylogenetic tree was visualized using ggplot and ggtree. In addition, resulting time scaled phylogeny can be viewed interactively and has been shared publicly on the NGS-SA NextStrain page at: https://nextstrain.org/groups/ngs-sa/COVID19-Alex-2021.04.29. Downstream analysis and visualization was performed using custom scripts in R^24^.

## Results

SARS-CoV-2 RT-PCR was positive at all time-points between day 0 and day 216, with mean Ct ranging from 16.4 to 31.6 (Table 1 & Fig. 1). Anti-S1 IgG and IgM were not detected at any time-point using a commercial rapid test kit. At study enrolment (day 6), HIV-1 RNA was 34,151 copies/mL and CD4+ cell count was 6 cells/µL. HIV-1 RNA remained elevated at all time points up to day 204, then decreased to <50 copies/mL two weeks following the switch to TLD. Until viral suppression, CD4+ cell count was persistently below 20 cells/µL. The antiretroviral drugs tenofovir, emtricitabine and efavirenz were detectable in plasma specimens only on day 71 and day 106. At enrolment, there were HIV-1 reverse transcriptase mutations associated with high-level resistance to efavirenz (V106M, K103R + V179D) and low-level resistance to tenofovir (K70Q). At the later-time points when antiretrovirals were detectable, the M184V mutation associated with high-level resistance to emtricitabine was also detected (Table S2).

**Table 1.** Results of SARS-CoV-2 PCR assays, antibody assays, HIV viral load and CD4+ count

| Time point | SARS-CoV-2 PCR | Mean Ct* | SARS-CoV-2 IgG | SARS-CoV-2 IgM | HIV-1 RNA (copies/mL) | CD4+ count (cells/μL) |
| --- | --- | --- | --- | --- | --- | --- |
| Day 0 | Positive | 18.5 | - | - | - | - |
| Day 6 | Positive† | 16.4† | Negative | Negative | 34 151 | 6 |
| Day 20 | Positive† | 19.2† | Negative | Negative | 6394 | 6 |
| Day 34 | Positive | 21.6 | Negative | Negative | 5477 | 5 |
| Day 71 | Positive | 19.2 | Negative | Negative | 4309 | 7 |
| Day 106 | Positive | 26.9 | Negative | Negative | 22 905 | 15 |
| Day 190 | Positive | 21.3 | Negative | Negative | 14 964 | 6 |
| Day 204 | Positive | 29.1 | Negative | Negative | <50 | 19 |
| Day 216 | Positive | 31.6 | Negative | Negative | - | 64 |
| Day 233 | Negative | >40 | Negative | Negative | <50 | 46 |

\* Mean Ct value calculated as mean of the Ct values for each of the three gene targets
† Specimens from day 6 and day 20 tested with the Taqpath™ COVID-19 RT-PCR assay (Thermo Fisher Scientific, Waltham, MA) targeting ORF1ab, N gene, and S gene; specimens from all other time points tested with the Allplex™ 2019-nCoV Assay (Seegene Inc., Seoul, South Korea) targeting E gene, N gene, and RdRp gene

**Figure 1.**
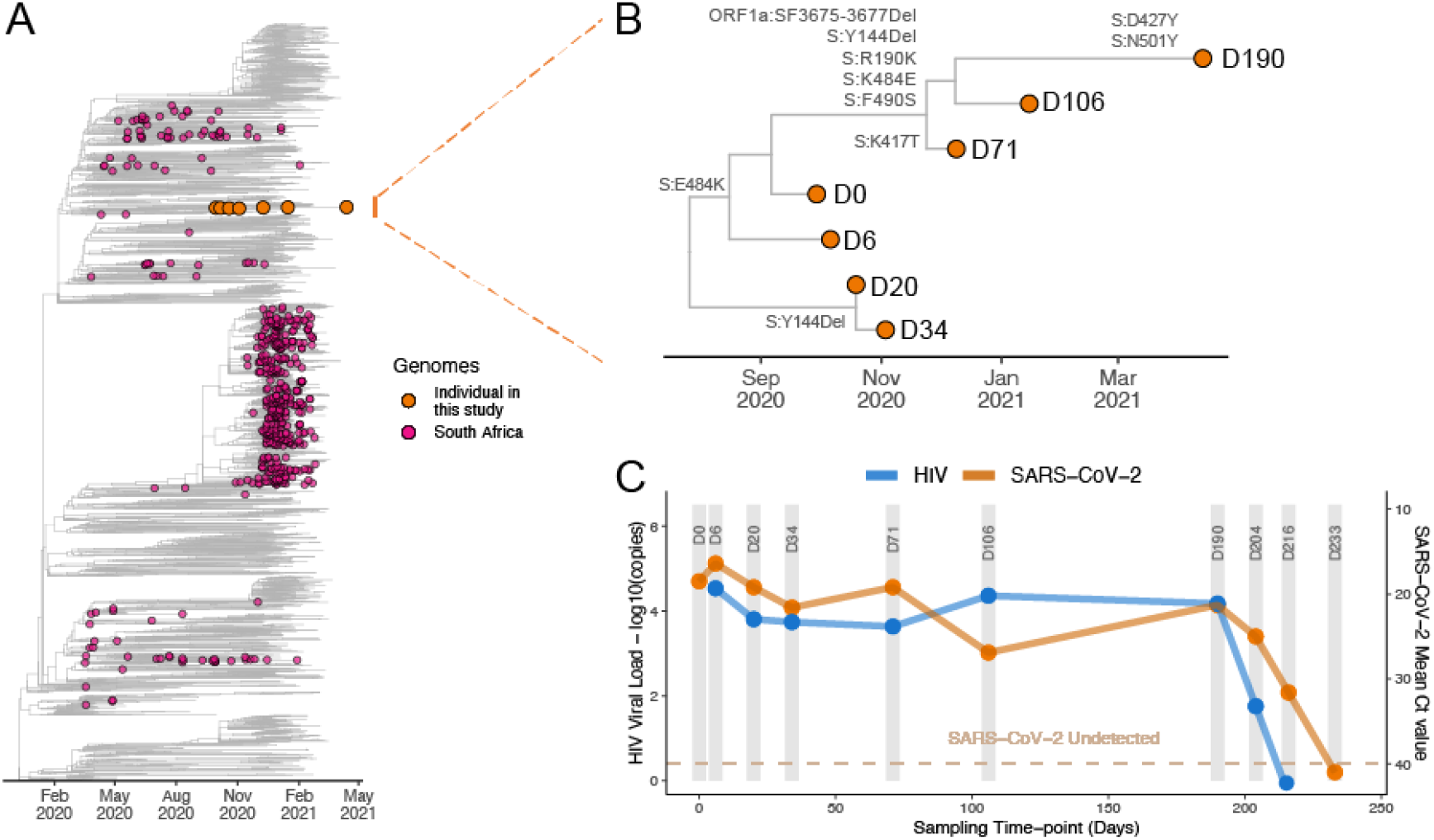
Phylogenetic analysis of seven SARS-CoV-2 whole genome sequences from a patient with advanced HIV. Shown in Panel A is a maximum-likelihood phylogenetic tree with patient sequences (orange) at seven time-points in relation to 486 representative South African and 3080 other global sequences. Panel B zooms in to show the relationship between the six patient-derived sequences from day 0, day 6, day 20, day 34, day 71, day 106 and day 190. Panel C shows the HIV-1 viral load in log scale (blue, left y-axis) and mean SARS-CoV-2 RT-PCR cycle threshold (Ct) value (orange, right y-axis) at each sampling time point.

We generated high quality whole SARS-CoV-2 genomes from samples at all seven time-points. Phylogenetic analysis was consistent with persistent infection and accelerated intra-host evolution (Fig. 1). All genomes fell within PANGO lineage B.1.1.273^25^, a South African lineage that was previously part of B.1.1.56, one of the most prevalent lineages in the first wave of the epidemic in the province of KwaZulu-Natal in South Africa, the location of the study^26^. The sequence at day 0 had the D614G mutation in addition to two additional spike mutations (G142V and D796Y). At day 6, the E484K substitution in the receptor-binding domain (RBD) emerged, along with a substitution in the S2 subunit (A1078V) (Fig. 2). The E484K mutation remained in the consensus sequence until day 34, but then persisted at low frequency up to day 190 (Suppl. Fig. 1, Table S3). At the later time points, there was a significant shift in the virus population, with the emergence at day 71 of the K417T and F490S mutations in the spike RBD, followed by the emergence of L455F and F456L at day 106. At day 190, there was the emergence of D427Y and N501Y.

**Figure 2.**
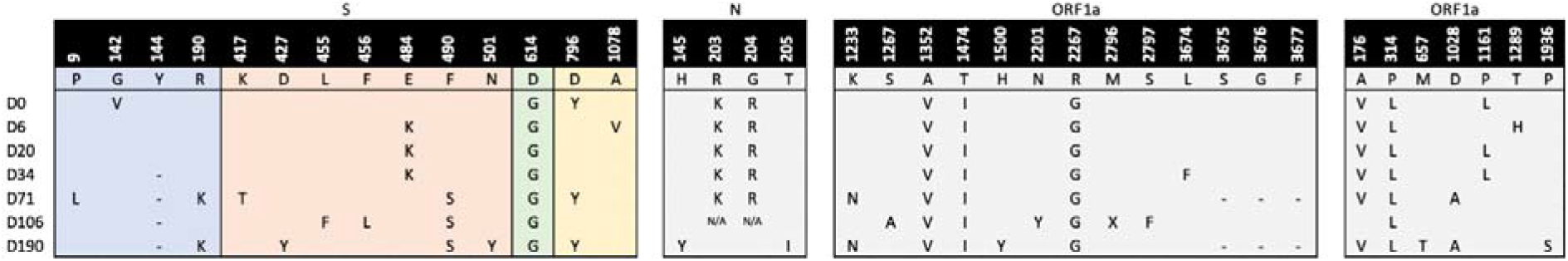
Locations of SARS-CoV-2 amino acid substitutions and deletions over time. Shown are the amino acid substitutions (marked by the substituted amino acid) and deletions (marked by a dash at the amino acid location) in the consensus sequences at each of the seven time-points (D0, D6, D20, D34, D71, D106 and D190). S spike, N nucleocapsid, ORF open reading frame.

## Discussion

We present a case of persistent SARS-CoV-2 infection with accelerated intra-host evolution in a patient with advanced HIV and antiretroviral treatment failure. Despite a short clinical illness of moderate severity, SARS-CoV-2 PCR positivity persisted up to 216 days. We demonstrate significant shifts in the virus population over that time, involving multiple mutations at key neutralizing antibody epitopes in the spike RBD and N terminal domain (NTD). Unlike many of the other reported cases, virus evolution was not driven by the receipt of immune-based therapies (convalescent plasma or monoclonal antibodies)^3-7,11^. Most other cases of persistent infection have been described in people with haematological malignancies or people receiving immunosuppressive therapies for solid organ transplants or other chronic medical conditions^3-10,12^. There is one other documented case of prolonged infection in a person with HIV -in that case the person was ART-naïve with profound immunosuppression and genome sequencing revealed only a single emergent mutation in spike (T719I) at day 53^12^. Although awaiting further viral and immunological analysis, the genomic and clinical data suggest that virus evolution may have been driven by selective pressure from an impaired neutralizing antibody response.

The E484K mutation in the spike RBD emerged very early in the course of infection, six days post-diagnosis and about 19 days post-symptom onset. This mutation is associated with resistance to class 2 neutralizing antibodies (NAbs)^27-33^. E484K has been observed in other cases of persistent infection, although in these cases it has emerged at a later stage in the course of infection^3,6,11^. At day 34, the Y144 deletion emerged and persisted up to day 106. This is in one of the recurrent deletion regions (RDR) in NTD that are associated with NAb escape^34^, and deletions in that specific region have been observed frequently in persistent infection^3,5-7,9-11,14^. As infection persisted, there were significant shifts in the virus population and the E484K mutation was replaced by mutations at other sites in the RBD associated with resistance to class 1, 2 and 3 NAbs (K417T and F490S at day 71 and then additionally L455F and F456L at day 106)^30^. The pattern of mutations is notable for RBD mutations at E484, K417 and N501, three of the signature mutations of the 501Y.V2 (B.1.351) variant of concern^15^. We believe this provides further evidence to support accelerated intra-host evolution as a plausible mechanism for the emergence of 501Y.V2 in South Africa.

There is some evidence that HIV is associated with an increased risk of more severe disease and death from COVID-19^17^, although in most studies the association is modest^36-39^. Two studies from South Africa have demonstrated that HIV is associated with suboptimal CD4+ T-cell and humoral immune responses to SARS-CoV-2, particularly in the absence of suppressive ART^17,40^; and one study from the United States suggested that HIV is associated with lower neutralizing antibody titres following natural infection^41^. In the case described here, based on the very low CD4+ count there was putative immunosuppression as a result of antiretroviral treatment failure and HIV drug resistance. The impairment of both cellular and humoral adaptive immunity from HIV was obviously profound enough to delay clearance of SARS-CoV-2^42^. Importantly, clearance of SARS-CoV-2 closely followed more effective suppression of HIV (Fig. 1C).

South Africa, has the largest HIV treatment programme in the world, with an estimated 5.2 million people on ART. Despite this, there remains a considerable burden of advanced HIV disease^43,44^. Disruptions to HIV programmes as a result of COVID-19 could potentially increase the burden of advanced HIV^45^. These findings highlight the importance of maintaining essential health services in high HIV prevalence settings, and accelerating progress towards 90-90-90 targets^46^. Preliminary results in people with well-controlled HIV suggest immune responses to COVID-19 vaccines are equivalent to those in HIV-negative people^47,48^, but more work is needed to understand immunogenicity and effectiveness in PLHIV and to design optimal dosing strategies, particularly for those with advanced HIV. If persistent infection does occur more frequently in the context of HIV, it may provide justification for prioritising people living with HIV for COVID-19 vaccination.

## Data Availability

All of the genomes are available at GISAID and short-read data at the Short Read Archive (SRA) of NCBI/EBI

