## Supplementary Material for "Persistent SARS-CoV-2 infection and intra-host evolution in association with advanced HIV infection"

**Table S1** Laboratory test results

| **Test** | **Normal range** | **Time point** | | | | |
| --- | --- | --- | --- | --- | --- | --- |
|  |  | **Day 0** | **Day 6** | **Day 71** | **Day 106** | **Day 190** |
| Haemoglobin | 12.0-15.0 g/dL | 12.9 | 11.9 | 12.9 | - | 11.6 |
| Platelets | 186-454 x 10^9^/L | 323 | 401 | 294 | - | 166 |
| White cell count | 3.90-12.60 x 10^9^/L | 9.60 | 7.10 | 4.11 | - | 2.30 |
| Lymphocyte count | 1.40-4.50 x 10^9^/L | 0.60 | - | 0.96 | - | 0.64 |
| Creatinine | 49-90 µmol/L | 43 | 61 | - | - | 49 |
| Bilirubin | 5-21 µmol/L | 4 | <2 | - | - | - |
| Alanine aminotransferase | 7-35 U/L | 24 | 44 | - | - | - |
| Alkaline phosphatase | 42-98 U/L | 96 | 76 | - | - | - |
| D-dimer | <0.25 mg/L | 0.36 | - | 0.65 | - | 0.32 |
| C-reactive protein | <10 mg/L | 113 | 22 | 40 | - | <10 |
| Sputum Xpert MTB/RIF Ultra |  | Negative | - | - | Negative | Negative |
| QuantiFERON-TB Gold Plus |  | Negative | Negative | Negative | Negative | Negative |
| Urine LAM assay |  | Negative | - | - | - | - |
| Cryptococcal antigen |  | Negative | - | - | - | Negative |

**Table S2** Results of HIV viral load, antiretroviral drug level and drug resistance assays

| **Time point** | **Antiretroviral therapy regimen** | **HIV RNA (copies/mL)** | **Antiretroviral drug levels (ng/mL)** | | |  | **Drug resistance mutations** | |
| --- | --- | --- | --- | --- | --- | --- | --- | --- |
|  |  |  | Tenofovir | Emtricitabine | Efavirenz |  | NRTI DRMs | NNRTI DRMs |
| Day 6 | TDF/FTC/EFV | 34 151 | - | - | - |  | K70KQ | K103R, V106M, V179D |
| Day 20 | TDF/FTC/EFV | 6394 | - | - | - |  | K70KQ | K103R, V106M, V179D |
| Day 34 | TDF/FTC/EFV | 5477 | - | - | - |  | K70KQ | K103R, V106M, V179D |
| Day 71 | TDF/FTC/EFV | 4309 | 19.6 | 305 | 1470 |  | K70KQ, M184V | K103R, V106M, V179D |
| Day 106 | TDF/FTC/EFV | 22 905 | <10 | 107 | 2460 |  | K70KQ, M184V | K103R, V106M, V179D |
| Day 190 | TDF/FTC/EFV | 14 964 | - | - | - |  | K70KQ | K103KNRS, V106VM, V179VD |
| Day 204 | TDF/3TC/DTG | 57 | - | - | - |  | - | - |

3TC, lamivudine; DRM, drug resistance mutation; EFV, efavirenz; FTC, emtricitabine; NNRTI, non-nucleoside reverse-transcriptase inhibitor; NRTI, nucleoside reverse-transcriptase inhibitor; TDF, tenofovir disoproxil fumarate

**Table S3** - Codon frequency table. This table shows the amino acid change, the nucleotide position of the genome, codon change and the frequency of the codon on the assembled genome.

| **Amino Acid change** | **Nucleotide change** | **Codon change** | **D0** | **D7** | **D20** | **D34** | **D71** | **D106** | **D190** |
| --- | --- | --- | --- | --- | --- | --- | --- | --- | --- |
| P9L | 21588C>T | 21587 CCA >CTA | CCA – 70  CTA - 0 | GAP | GAP | CCA – 7  CTA – 0 | CCA – 94  CTA – 613 | GAP | CCA – 0  CTA – 7 |
| T95I | 21846C>T | 21845 ACT > ATT | ACT – 146  ATT – 0 | ACT – 6105  ATT - 9 | ACT – 2667  ATT - 298 | ACT – 935  ATT – 1626 | ACT – 1379  ATT – 36 | ACT – 11843  ATT – 576 | ACT – 231  ATT – 10 |
| G142V | 21987G>T | 21986 GGT>GTT | GGT – 10  GTT – 40 | GGT – 1386  GTT - 622 | GGT – 880  GTT - 313 | GGT – 954  GTT – 96 | GGT – 480  GTT – 5 | GGT – 3621  GTT – 12 | GGT – 159  GTT – 24 |
| R190K | 22131G>A | 22130  AGG>AAG | AGG – 46  AAG – 0 | AGG – 5591  AAG – 8 | AGG – 1702  AAG – 1 | AGG – 1685  AAG - 2 | AGG – 222  AAG – 1096 | GAP | AGG – 16  AAG – 41 |
| S341S | 22675C>T | 22673  TCC>TCT | TCC – 0  TCT – 77 | TCC – 6  TCT – 3300 | TCC – 0  TCT – 17 | TCC – 0  TCT – 7 | TCC – 0  TCT – 3 | TCC – 6  TCT – 0 | GAP |
| K417T | 22812A>G | 22811 AAG>ACG | AAG – 117  ACG – 0 | AAG – 3237  ACG - 1 | AAG – 3209  ACG – 0 | AAG – 2587  ACG – 1 | AAG – 299  ACG – 1362 | AAG – 32  ACG – 2 | GAP |
| K417N | 22813G>T | 22811 AAG>AAT | AAG – 117  AAT – 0 | AAG – 3237  AAT - 6 | AAG – 3209  AAT – 313 | AAG – 2587  AAT –  21 | AAG – 299  AAT – 5 | AAG – 32  AAT – 4 | GAP |
| D427Y | 22841G>T | 22841 GAT>TAT | GAT – 107  TAT – 0 | GAT – 3856  TAT – 3 | GAT – 4197  TAT – 0 | GAT – 3433  TAT - 0 | GAT – 4198  TAT – 0 | GAT – 221  TAT – 20 | GAT – 19  TAT - 103 |
| L455F | 22927G>T | 22925 TTG>TTT | TTG – 11  TTT – 0 | TTG – 3713  TTT – 9 | TTG – 3219  TTT – 0 | TTG – 4331  TTT – 2 | TTG – 2983  TTT – 4 | TTG –527  TTT – 317 | TTG – 114  TTT – 0 |
| F456L | 22928T>C | 22928  TTT>CTT | TTT – 12  CTT – 0 | TTT – 3739  CTT – 6 | TTT – 3248  CTT – 0 | TTT – 4361  CTT – 0 | TTT – 2971  CTT – 3 | TTT – 537  CTT – 310 | TTT – 139  CTT – 2 |
| A475V | 22986C>T | 22985  GCC>GTC | GCC – 17  GTC – 0 | GCC – 3409  GTC - 11 | GCC – 3629  GTC – 1218 | GCC – 2874  GTC – 2421 | GCC – 3396  GTC – 101 | GCC – 901  GTC – 23 | GCC – 177  GTC – 14 |
| E484K | 23012G>A | 23012  GAA>AAA | GAA – 17  AAA – 0 | GAA – 56  AAA - 2063 | GAA – 1632  AAA – 2881 | GAA – 2395  AAA – 2139 | GAA – 3147  AAA – 196 | GAA – 688  AAA – 40 | GAA – 180  AAA – 54 |
| F490S | 23031T>C | 23030  TTT>TCT | TTT – 18  TCT – 0 | TTT- 1605  TCT - 2 | TTT – 4067  TCT – 0 | TTT – 4007  TCT – 8 | TTT – 341  TCT – 2912 | TTT – 182  TCT –  446 | TTT – 89  TCT – 125 |
| N501Y | 23063A>T | 23063 AAT>TAT | AAT – 13  TAT – 0 | AAT – 810  TAT – 2 | AAT – 2567  TAT – 358 | AAT – 2856  TAT – 29 | AAT – 2511  TAT – 14 | AAT – 282  TAT – 13 | AAT – 20  TAT – 180 |
| L518L | 23114C>T | 23114  CTA>TTA | CTA – 0  TTA – 8 | CTA – 0  TTA - 584 | CTA – 302  TTA – 10 | CTA – 304  TTA – 56 | CTA – 157  TTA – 1166 | CTA – 0  TTA – 240 | CTA – 93  TTA – 85 |
| D796Y | 23948G>T | 23948  GAT>TAT | GAT – 16  TAT – 70 | GAT – 4293  TAT – 16 | GAT – 1235  TAT – 477 | GAT – 568  TAT – 649 | GAT – 97  TAT – 791 | GAT – 7  TAT – 0 | GAT – 20  TAT –  89 |
| A1078V | 24795C>T | 24974  GCT>GTT | GCT – 219  GTT – 3 | GCT – 772  GTT - 4833 | GCT – 4397  GTT – 1 | GCT – 2519  GTT – 12 | GCT – 1462  GTT – 30 | GCT – 13695  GTT – 74 | GCT – 310  GTT – 17 |

**Figure S1** – Frequency of nucleotide positions at Spike and effects on amino acid mutations.
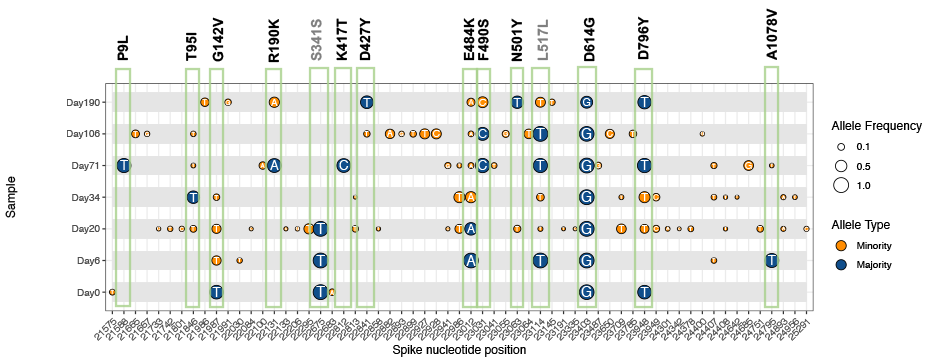
